# Development and Implementation of Dried Blood Spot-based COVID-19 Serological Assays for Epidemiologic Studies

**DOI:** 10.1101/2021.11.25.21266786

**Authors:** Marcus P Wong, Michelle A Meas, Cameron Adams, Samantha Hernandez, Valerie Green, Magelda Montoya, Brett M Hirsch, Mary Horton, Hong L Quach, Diana L Quach, Xiaorong Shao, Indro Fedrigo, Alexandria Zermeno, Julia Huffaker, Raymond Montes, Alicia Madden, Sherri Cyrus, David McDowell, Phillip Williamson, Paul Contestable, Mars Stone, Josefina Coloma, Michael P Busch, Lisa F Barcellos, Eva Harris

## Abstract

Serological surveillance studies of infectious diseases provide population-level estimates of infection and antibody prevalence, generating crucial insight into population-level immunity, risk factors leading to infection, and effectiveness of public health measures. These studies traditionally rely on detection of pathogen-specific antibodies in samples derived from venipuncture, an expensive and logistically challenging aspect of serological surveillance. During the COVID-19 pandemic, guidelines implemented to prevent the spread of SARS-CoV-2 infection made collection of venous blood logistically difficult at a time when SARS-CoV-2 serosurveillance was urgently needed. Dried blood spots (DBS) have generated interest as an alternative to venous blood for SARS-CoV-2 serological applications due to their stability, low cost, and ease of collection; DBS samples can be self-generated via fingerprick by community members and mailed at ambient temperatures. Here, we detail the development of four DBS-based SARS-CoV-2 serological methods and demonstrate their implementation in a large serological survey of community members from 12 cities in the East Bay region of the San Francisco metropolitan area using at-home DBS collection. We find that DBS perform similarly to plasma/serum in enzyme-linked immunosorbent assays and commercial SARS-CoV-2 serological assays. In addition, we show that DBS samples can reliably detect antibody responses months post-infection and track antibody kinetics after vaccination. Implementation of DBS enabled collection of valuable serological data from our study population to investigate changes in seroprevalence over an eight-month period. Our work makes a strong argument for the implementation of DBS in serological studies, not just for SARS-CoV-2, but any situation where phlebotomy is inaccessible.

## Introduction

Serological surveillance of infectious diseases provides estimates of the incidence and prevalence of infections, generating actionable public health knowledge – such as which populations are disproportionately affected by an infectious disease outbreak or the effectiveness of public health measures in curbing infections. SARS-CoV-2, the causative agent of the COVID-19 pandemic, is responsible for at least 250 million infections and 5 million deaths since its emergence in late 2019 to date (1). Given that approximately 35% of all SARS-CoV-2 infections are asymptomatic (2, 3), with proportions varying substantially by age, serological surveillance studies are a critical tool to enable estimation of the incidence of recent infection and prevalence of past infections at the population level. Most serological studies rely on venous blood from study participants to derive plasma or serum and therefore require an invasive phlebotomy procedure and an intact cold chain for collection, shipment, and storage to ensure sample stability prior to testing. However, during the COVID-19 pandemic, implementation of public health guidelines to curb the spread of SARS-CoV-2, such as lockdowns and social distancing, made the collection of blood to obtain serum or plasma for serosurveillance studies logistically challenging, especially for sizeable community-based studies focused on a large geographic region.

Dried blood spots (DBS) are an alternative blood collection method for serological testing and have been used extensively in screening for other viruses, including Hepatitis B, HIV, and Ebola in both clinical and non-clinical settings (4–6). DBS have advantages over traditional venous blood sampling (7): DBS are stable at room temperature for extended periods of time and can be easily shipped by mail and stored (8), DBS require considerably less blood than traditional phlebotomy, and DBS can be collected by study participants themselves via fingerprick. These qualities make DBS an attractive alternative to venous blood collection, especially when traditional phlebotomy is inaccessible, such as in resource-poor settings or because of pandemic-related public health restrictions.

There has been interest in developing DBS-based serological methods for SARS-CoV-2 serosurveillance. Several studies have investigated the feasibility and performance of DBS-based serological assays, including enzyme-linked immunosorbent assays for anti-spike (S) and receptor binding domain (RBD) antibodies (9–13), as well as multiplexed assay formats for simultaneous detection of spike, RBD, and nucleocapsid (N) antibodies (14). Commercial assays on automated platforms like the Roche Elecsys, which can detect anti-N or anti-S antibodies, have also been evaluated for use with DBS samples. These assays have the advantage of high-throughput processing and lower personnel requirements (15, 16). Based on testing paired plasma/serum and DBS samples, these studies demonstrated robust agreement between the sample types. In addition, several studies have shown that serological assays performed on DBS samples prepared by study participants in the home and mailed to labs for processing can reliably detect SARS-Cov-2 antibodies, providing evidence that DBS are a promising alternative to plasma or serum for SARS-CoV-2 serosurveillance (13–15, 17). However, implementation of DBS-adapted serological methods for large community-based serological surveillance has not been widely demonstrated for tracking immune responses due to natural infections and COVID-19 vaccines.

In this study, we present the validation of four DBS-based serological assays against SARS-CoV- 2 S and N antibodies and detail their implementation in a large, serological survey with at-home sample collection, as well as in ancillary longitudinal vaccine studies. We assessed the performance of DBS-based lab-developed anti-S and anti-N IgG ELISAs and two commercial assays (the Ortho anti-S and Roche anti-N Total Ig assays) and evaluated their ability to detect long-term antibody responses and vaccine-induced antibody kinetics. Using these assays in a serial testing algorithm, we analyzed a total of 14,782 DBS samples from a longitudinal cohort of individuals living in 12 cities in the East Bay region of the San Francisco metropolitan area at 3 timepoints between July 2020 and April 2021 for antibodies against SARS-CoV-2 S and N. Results from this study demonstrate that DBS are a practical sampling method for serology-based epidemiology studies, enabling in-home self-sampling for serosurveillance in situations where traditional phlebotomy is inaccessible, impractical, or too costly.

## Methods

### Materials

We obtained DBS cards (Tropbio Filter Paper Blood Collection Disks) from Cellabs. Reagents for the DBS Elution Buffer (63mM K_2_HP0_4_, 28mM KH_2_PO_4_, 139mM NaCl, 5g/L Sodium Azide, 5g/L Caesin, 50g/L Probumin BSA in H_2_0) were obtained from Sigma-Aldrich. SARS-CoV-2 soluble trimeric S and N proteins used in ELISAs developed at UC Berkeley were provided by Dr. John Pak (Chan-Zuckerberg Biohub) and Dr. Aubree Gordon (University of Michigan), respectively.

### Human Subjects Ethics Statement

Samples for DBS validation studies were collected from participants consented under IRB #11- 06262 approved by the University of California, San Francisco Committee on Protection of Human Subjects. Use of samples from convalescent plasma donors but did not involve human subjects based on anonymization of data and routine consent for blood donation testing that includes use of residual samples for research purposes. Samples for validation of the S and N IgG ELISAs were obtained from Dr. Benjamin Pinsky (Stanford University) and Dr. Bryan Greenhouse (UC San Francisco) and were pre-collected and de-identified.

All participants in the East Bay COVID study provided informed consent for the initial screening phase of the study. All those participating in the sampling phase of the study provided their informed consent for each sample/data collection round. The study was approved by the University of California, Berkeley Committee on Protection of Human Subjects (Protocol #2020-03-13121).

### Validation Studies

Plasma and DBS samples were collected from COVID-19 convalescent patients and SARS-CoV- 2-infected individuals. Reconstituted DBS were generated by mixing plasma with anticoagulated, plasma-depleted whole blood and spotted on DBS cards. Four sets of samples were generated for validation studies. The first sample set consisted of 39 paired plasma and reconstituted DBS samples from previously SARS-CoV-2-infected individuals and 100 paired samples from individuals without previous SARS-CoV-2 infection. The second sample set consisted of paired plasma and reconstituted DBS from 10 COVID-19 convalescent plasma donors sampled longitudinally between 0 and 246 days from their first donation. The third set of samples consisted of paired plasma and fingerstick DBS samples generated from 12 vaccinated individuals sampled before their first dose, after their first dose, and after their second dose. Four other vaccinated individuals were sampled by fingerstick DBS over a period of 210 days, after their first, second, and third doses. The plasma-based S and N IgG ELISAs IgG ELISA were validated using convalescent plasma samples collected >8 days post-symptom onset from 60 hospitalized, PCR-confirmed severe COVID-19 cases, 57 mild or subclinical cases, and samples collected before 2020 from 131 unexposed persons as described previously (18).

### East Bay COVID (EBCOVID) Study Design

Recruitment and selection of study participants was completed in a screening phase followed by a longitudinal sampling phase with three timepoints or “rounds” (Figure 1). In the screening phase, all residential addresses within the East Bay cities and communities of Albany, Berkeley, El Cerrito, El Sobrante, Emeryville, Hercules, Kensington, Oakland, Piedmont, Pinole, Richmond, and San Pablo (∼307,000 residential households) were mailed an invitation to participate. The household member aged 18 or older with the next birthday was invited to complete a consent form and screening questionnaire. Spanish versions of study invitations and all study materials were also utilized.

**Figure 1.**
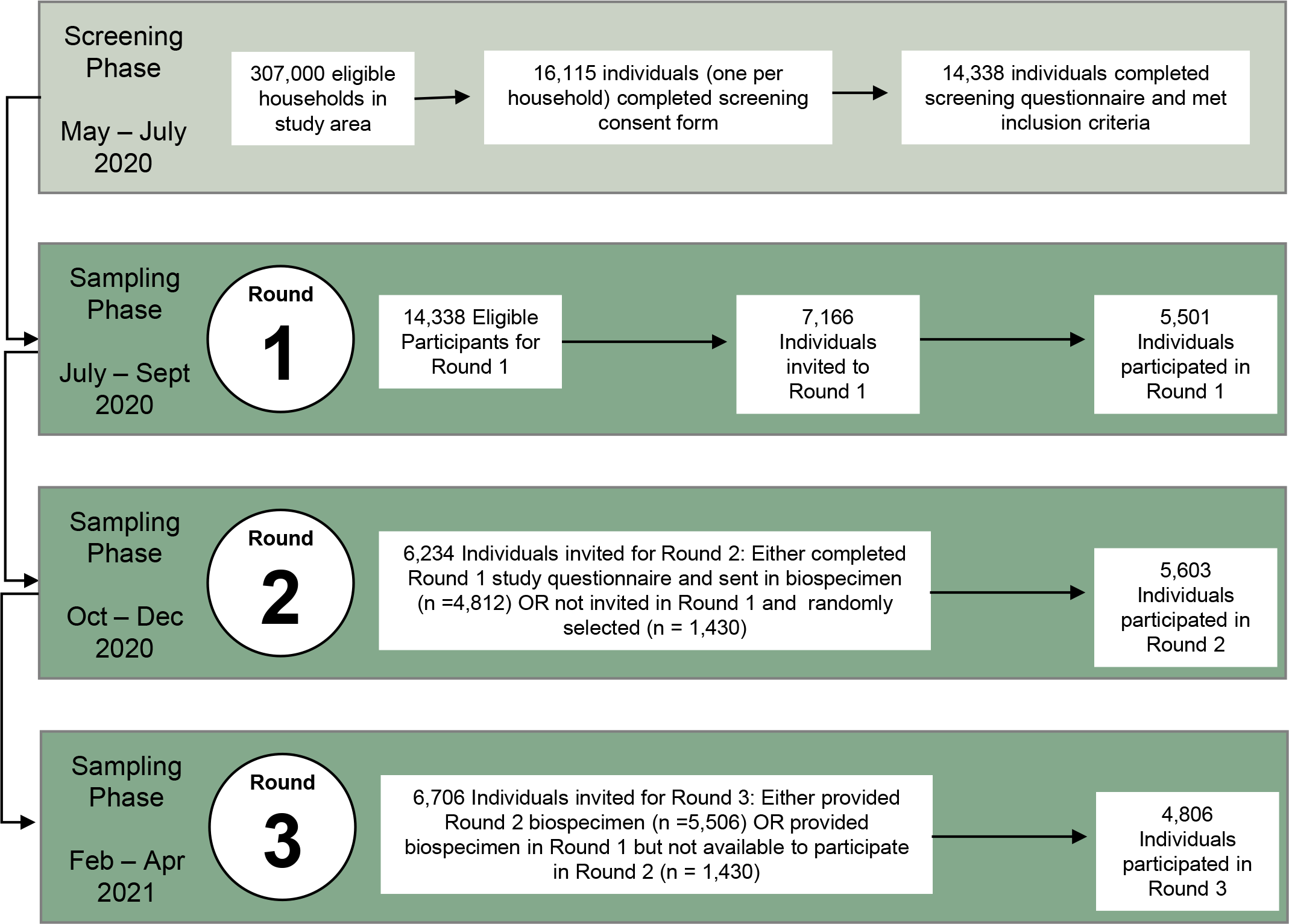
Schematic of the East Bay COVID (EBCOVID) study. There were 2 phases to the study: (A) the screening phase, where participants were recruited and screened for eligibility for inclusion into our study, and (B-D) the sampling phase, where study participants were invited to provide biospecimens, including DBS, in 3 separate rounds.

Of the 16,115 residents who consented and completed the screening procedures between May-July 2020, 1,777 individuals did not meet the inclusion criteria and were excluded (Figure 1). Eligible participants were required to be the household member with the next birthday, live within the study region, be willing to provide biospecimens (including DBS) and questionnaire responses, read and speak English or Spanish, and have internet access and a valid email address.

The target sample size for the sampling phase was 5,500 participants. To obtain a sample that resembled the racial and ethnic proportions reported in the 2018 American Community Survey (ACS) for the study region, we ranked screening participants for study inclusion. Black and/or Hispanic individuals were ranked the highest (n=1,556) followed by other non-White individuals (n=1,939). White individuals were randomly ranked to complete the remaining participant slots. Individuals ranked between 1 and 5,500 were offered study enrollment, and non-respondents were replaced with the next highest ranked individuals who had not yet been offered study entry.

Biospecimens and questionnaire data were collected during three sampling rounds. Approximate dates for each round were July-September 2020, October-December 2020, and February-April 2021. For each round of data and biospecimen collection, individuals who had participated in the previous round were contacted to confirm their willingness to participate in the next round. If participation was declined, individuals from the pool of screening participants who had not yet participated in a sampling round were invited as needed. This resulted in 5,501 participants in Round 1, 5,603 participants in Round 2, and 4,806 participants in Round 3 (Figure 1). This corresponds to participation rates of 76.8%, 89.8%, and 87.3% across the study rounds, respectively. A biospecimen collection kit was developed and assembled by investigators and sent to each participant via Federal Express (FedEx). Both written instructions and an instructional video for at-home sample collection, including fingerstick blood sampling and preparation of DBS, were provided to all study participants. Both English and Spanish versions were made available.

### Quality Control of DBS

DBS received from participants were assessed for quality. We evaluated each DBS on a scale from 0 to 3, with 0 representing a blank DBS, 1 representing an incompletely filled DBS on one side, 2 representing a DBS card saturated fully on only one side, and 3 representing a DBS fully saturated on both sides. Participants who provided at least two DBS samples with a cumulative score of 2 or higher were processed in our serological assays; samples that did not meet this criterion were labelled as “Quality Not Sufficient” (QNS).

### Reconstitution of Blood Spots

Two DBS were removed from the Tropbio disk and placed in a 2-mL screwcap tube. Five hundred uL of elution buffer was added into the tube and vortexed for 20 seconds. If DBS numbers were limited, one spot was eluted in 250uL of elution buffer instead. DBS were incubated in elution buffer overnight at 4°C, then spun down at 10,600xg for 10 minutes (min) at 4°C. The DBS eluate was then transferred to a fresh tube and stored at 4°C until analysis.

### ELISA (N and S)

DBS eluates and plasma were evaluated for the presence of IgG against SARS-CoV-2 S and NC using an in-house direct ELISA as previously described(18). Briefly, SARS-CoV-2 antigens were coated in 96-well Nunc Maxisorp ELISA plates (ThermoFisher) overnight at 4°C. Plates were then blocked in 2.5% non-fat dry milk in PBS for 2 hours (h) at 37°C. Plates were washed 3X with PBS (Gibco), and 100uL of DBS eluate or plasma was diluted 1:100 in 1% non-fat dry milk and incubated at 37°C for 1h. Plates were then washed 5 times with 0.05% PBS-Tween-20, and 100uL of goat-α-human IgG horseradish peroxidase (HRP) secondary antibody (Fisher) diluted in 1% non-fat dry milk was added. After incubating for 1h at 37°C, plates were washed 5 times in 0.05% PBS-Tween-20 and once in PBS. Wells were developed with TMB (3,3’,5,5’- Tetramethylbenzidine, ThermoFisher) for exactly 5 minutes, and the reaction was stopped with 2M H_2_SO_4._ Plates were read on a plate reader at 490nm. Endpoint-titer ELISAs were performed as above, except that the DBS eluate was serially diluted 1:4 or 1:5 eight times in DBS eluate before addition to the plate. Endpoint titers were calculated using the five-parameter logistic equation function in GraphPad Prism 8, using 0.34 as the negative cut-off.

### Commercial Assays (Ortho S & Roche N Total IgG Assays)

DBS eluates prepared as above (or paired plasma samples for assay validation) were tested with the Ortho VITROS_®_ Anti-SARS-CoV-2 S1 Total Ig assay (https://www.fda.gov/media/136967/download) or the Roche Elecsys ® Anti-SARS-CoV-2 N Total Ig assay (https://www.fda.gov/media/137605/download) according to the manufacturers’ instructions. Briefly, the Ortho VITROS Anti-SARS-CoV-2 total (CoV2T, Ortho-Clinical Diagnostics, Inc.) was used to detect total (IgG, IgM, and IgA) antibodies to SARS-CoV-2 Spike S1 protein. DBS eluates or plasma samples were loaded on Ortho VITROS XT-7600 or 3600 instruments (Ortho-Clinical Diagnostics, Inc.) and programmed for the CoV2T test following the manufacturer’s instructions. The S1 antigens coated on the assay wells bind anti-S1 antibodies from human serum which, in turn, bind to a secondary HRP-labeled S1 antigen in the conjugate reagent, forming a sandwich. The addition of signal reagent containing luminol generates a chemiluminescence reaction that is measured by the system and quantified as the ratio of the signal relative to the cut-off value (S/Co) generated during calibration. A S/Co ≥1 was considered positive in plasma, while a S/Co ≥0.7 in DBS eluate was considered indeterminate or grey zone (see Results).

The Roche Elecsys Anti-SARS-CoV-2 immunoassay (Roche N) was processed on the Cobas e441 analyzer (Roche Diagnostics) to detect total antibodies against the SARS-CoV-2 N protein. DBS eluates or plasma samples are initially incubated with biotinylated and ruthenium-labeled SARS-CoV-2 recombinant N antigens, and any anti-N antibody present in the solution is sandwiched between the two. Subsequently, streptavidin-coated microparticles are added to the mixture to bind the biotin. The magnetic particles drive the complexes to the electrode, where a chemiluminescent signal is emitted and measured as the ratio between the signal and the cut-off obtained during calibration. A S/Co≥1 in plasma was considered positive, while a S/Co ≥0.045 in DBS eluate was considered positive (see Results).

### Statistics

Statistical analysis was performed using GraphPad Prism Version 8 (GraphPad Software) and the R statistical programming package.

All materials and data are available upon request

## Results

### DBS are a viable replacement for plasma in multiple serological assay formats

To evaluate whether DBS samples could be a viable replacement for plasma in serosurveillance studies, we first validated the performance of DBS eluates on both an in-house ELISA detecting anti-S IgG and the Ortho CoV2T. To generate DBS for the validation study, we mixed COVID convalescent plasma (CCP) samples from 39 RT-PCR-confirmed SARS-CoV-2-positive patients and 100 pre-2020 negative controls with anticoagulated plasma depleted whole blood to reconstitute whole blood and applied the mixture to the DBS cards (∼15 ul per DBS). Eluates from these reconstituted DBS and their paired plasma samples were then tested by both serological assays.

We first tested DBS eluates in our previously validated anti-S IgG ELISA (18). We determined a new cut-off for the anti-S IgG ELISA using DBS eluate as the input (DBS ELISA) as an optical density (OD_450)_ value of 0.34 via receiver-operator curve (ROC) analysis using the convalescent samples following PCR diagnoses and the pre-2020 samples as the reference standards. This cut- off resulted in an overall sensitivity of 97.62% and specificity of 100% for the ELISA, which demonstrated 100% concordance in the SARS-CoV-2-positive samples between the reconstituted DBS eluate and plasma (Figure 2A-B, S1). When the recommended threshold (S/Co <u>≥</u>1) on the Ortho CoV2T for detection of anti-S antibodies in plasma/serum was used for DBS eluates, ROC analysis yielded a sensitivity of 79.5% and specificity of 100%, with an 80% concordance in the SARS-CoV-2-positive samples between the reconstituted DBS eluates and plasma (Figure 2C-D). Both assays showed a linear relationship between plasma and DBS values (ELISA, r^2^=0.93; Vitros r^2^=0.78; Figure 2 A,C).

**Figure 2.**
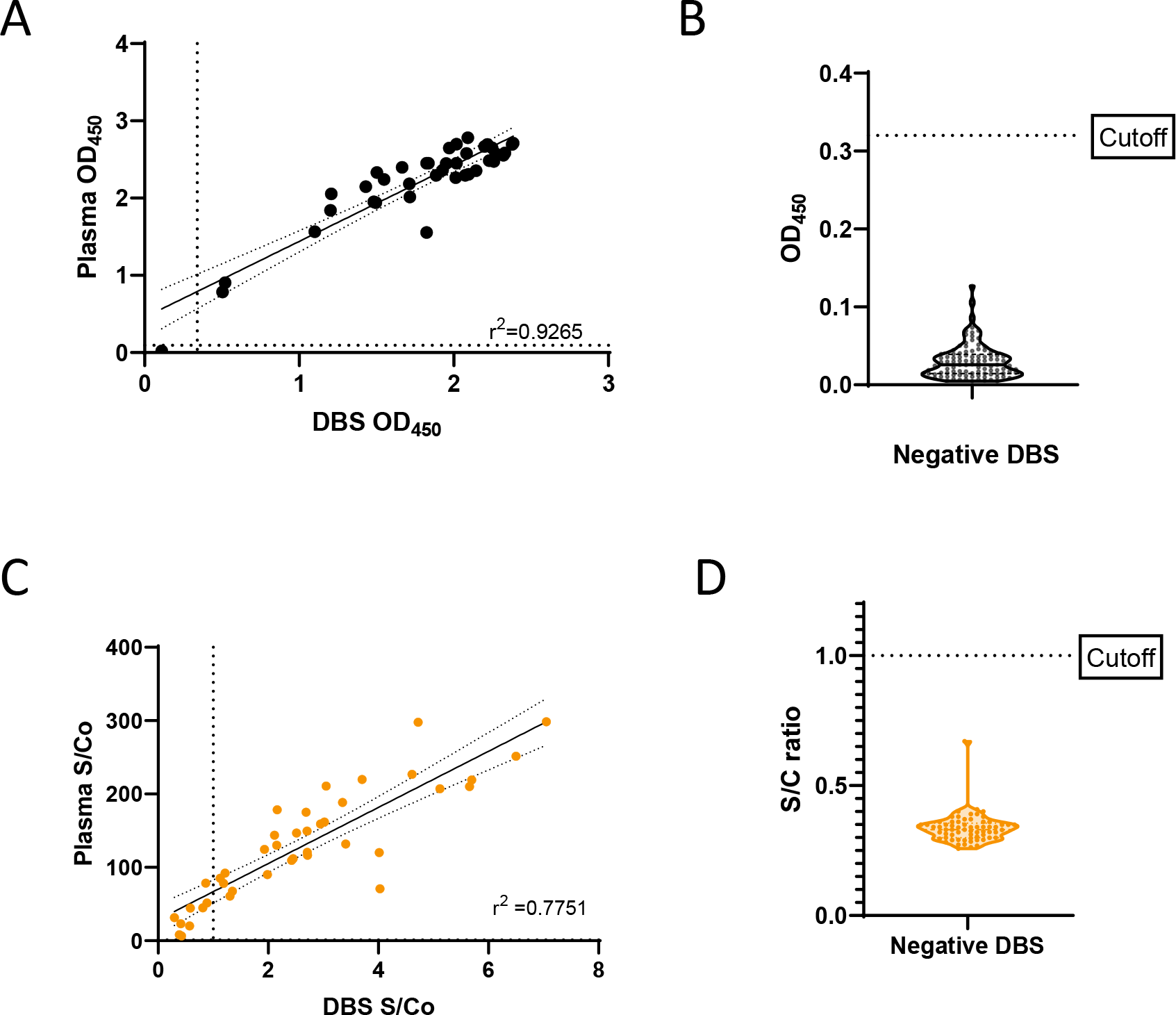
Validation of DBS in anti-S serological assays. Paired DBS and plasma samples (n=39) from previously SARS-CoV-2-infected individuals were compared in (A) the anti-S IgG ELISA and (C) the Ortho COV2T assay. DBS samples (n=100) from individuals without previous SARS-CoV-2 infection were analyzed by (B) the anti-S IgG ELISA and (D) the Ortho COV2T assay. Cut-offs for the assays are denoted by dashed lines. Linear regression comparing IgG levels between sample types (A and C) is depicted by a solid line with 95% confidence intervals (CI).

We found that the reduced sensitivity of the Ortho CoV2T assay could be explained by the diluted nature of the DBS eluate, as compared to the Ortho CoV2T assay using undiluted plasma. We found that diluting plasma 1:40 gave similar values as the DBS eluates (Figure S2). In contrast, the in-house ELISA was developed using a 1:100 dilution of plasma; thus, DBS eluates performed similarly to plasma in this assay format.

### Validation of anti-N serological assays

The introduction of S-based vaccines during the study period necessitated the implementation of N-based serology testing to distinguish between vaccinated and naturally infected individuals in the EBCOVID study. Therefore, we validated an anti-N IgG ELISA for both DBS and serum as well as the Roche N assay for use with DBS eluates. We determined the positivity cut-off for the anti-N IgG ELISA as an OD_490_ value of 0.32 via ROC analysis. This cutoff resulted in a sensitivity of 97.1% and specificity of 91.5% when testing serum samples (Figure S3A).

We performed validation of DBS eluates for the N DBS ELISA using the reconstituted DBS sample set used for the validation of our anti-S antibody assays and determined a positivity cut- off for DBS eluates as an OD_490_ value of 0.32 via ROC analysis (Figure S3B). This cut-off gave the ELISA an overall sensitivity of 89.7% and specificity of 90.4% and 87.1% concordance in the SARS-CoV-2 positive samples between the reconstituted DBS eluate and plasma (Figure 3A-B). Overall, the DBS eluates and plasma performed similarly in the anti-N IgG ELISA, comparable to our results for the anti-S IgG ELISA.

**Figure 3.**
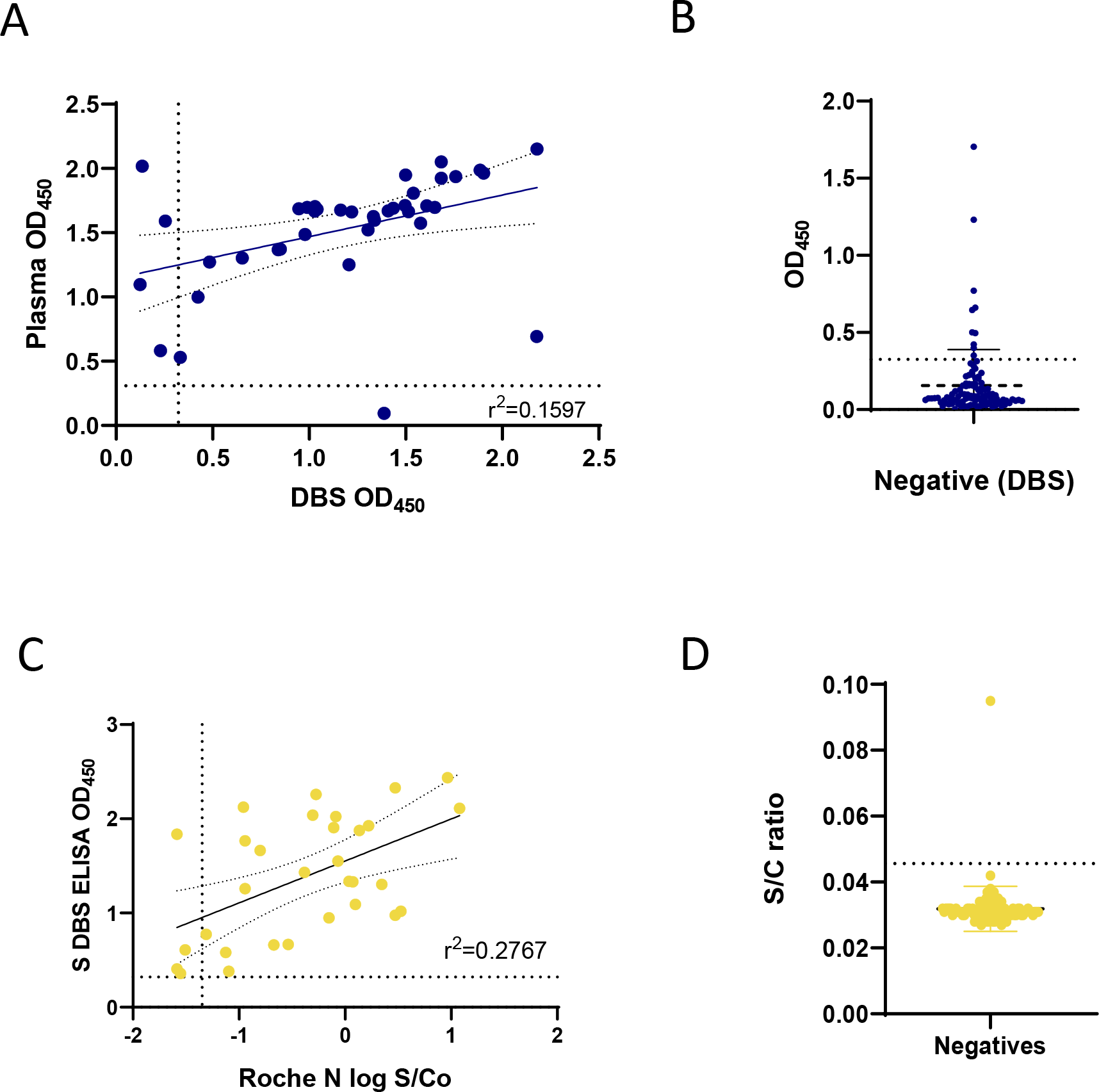
Validation of DBS in anti-N serological methods. Paired DBS and plasma samples (n=39) from previously SARS-CoV-2-infected individuals were compared in (A) the anti-N IgG ELISA. DBS samples (n=100) from individuals without previous SARS-CoV-2 infection were analyzed by (B) the anti-N IgG ELISA. Validation of DBS on the Roche N assay was performed on DBS samples derived from study participants in Round 2 of the EBCOVID study. (C) Samples considered SARS-CoV-2-seropositive (n = 33) from the anti-S IgG DBS ELISA were analyzed by the Roche N assay. (D) Samples considered SARS-CoV-2-seronegative (n=99) by the Ortho COV2T assay were analyzed by the Roche N assay. Cut-offs for the assays are denoted by dashed lines. Linear regression comparing IgG levels between sample types is depicted by a solid line with 95% CI.

Due to limited sample availability from our validation sample set, we validated the use of DBS eluates on the Roche N assay using reserved eluates from our Round 2 sampling that preceded vaccine approvals and rollout (Figure 1C). Using our anti-S IgG DBS ELISA as the reference standard, we determined a new positivity cut-off for the Roche N assay (S/Co ≥0.045). With this cut-off, the Roche N assay displayed a sensitivity of 86.7% and specificity of 97.9% using the DBS format (Figure 3C-D).

### DBS enables serosurveillance in a large longitudinal study during a pandemic

Our validation studies demonstrated that DBS are a reliable replacement for plasma, and thus we implemented the use of DBS cards as part of the EBCOVID study. The DBS collected from participants were assessed for quality before processing. Only samples with adequate numbers of saturated DBS were processed in our serological assays; samples that did not meet this criterion were labelled as “Quality Not Sufficient” (QNS), as described above. Overall, we tested 14,782 qualified DBS samples from study participants over three rounds of testing.

In our first round of testing (July to September 2020), we analyzed 4,670 DBS samples using the Ortho CoV2T Assay and detected anti-S antibodies in 29 individuals as positive (S/Co≥1.0), giving an unweighted seroprevalence of 0.6%. Analysis of Round 1 results revealed that 1.08% of samples had S/Co ratios between 0.7 and 1.5, close to the positivity cutoff of 1.0 set by the manufacturer (Figure S4). Given the linear relationship of DBS eluate to plasma and the dilutional effect attributable to DBS elution seen in our validation of the Ortho CoV2T assay, we reasoned that these samples were likely antibody-positive samples with lower seroreactivity. To better resolve samples with grey-zone reactivity in the Ortho CoV2T in subsequent rounds, we devised an algorithm utilizing the in-house S DBS ELISA to retest samples with Ortho CoV2T S/Co results that fell between 0.7 and 2.0.

We implemented this algorithm in Round 2 (October to December 2020), testing 5,308 samples by the Ortho Total Ig assay. Of these, 317 samples with S/Co values >0.7 and <2.0 were reflexed to the anti-S IgG ELISA, resulting in 33 S antibody-positive samples, giving an unweighted seroprevalence of 0.6% in the cohort in Round 2 (Figure 4).

**Figure 4.**
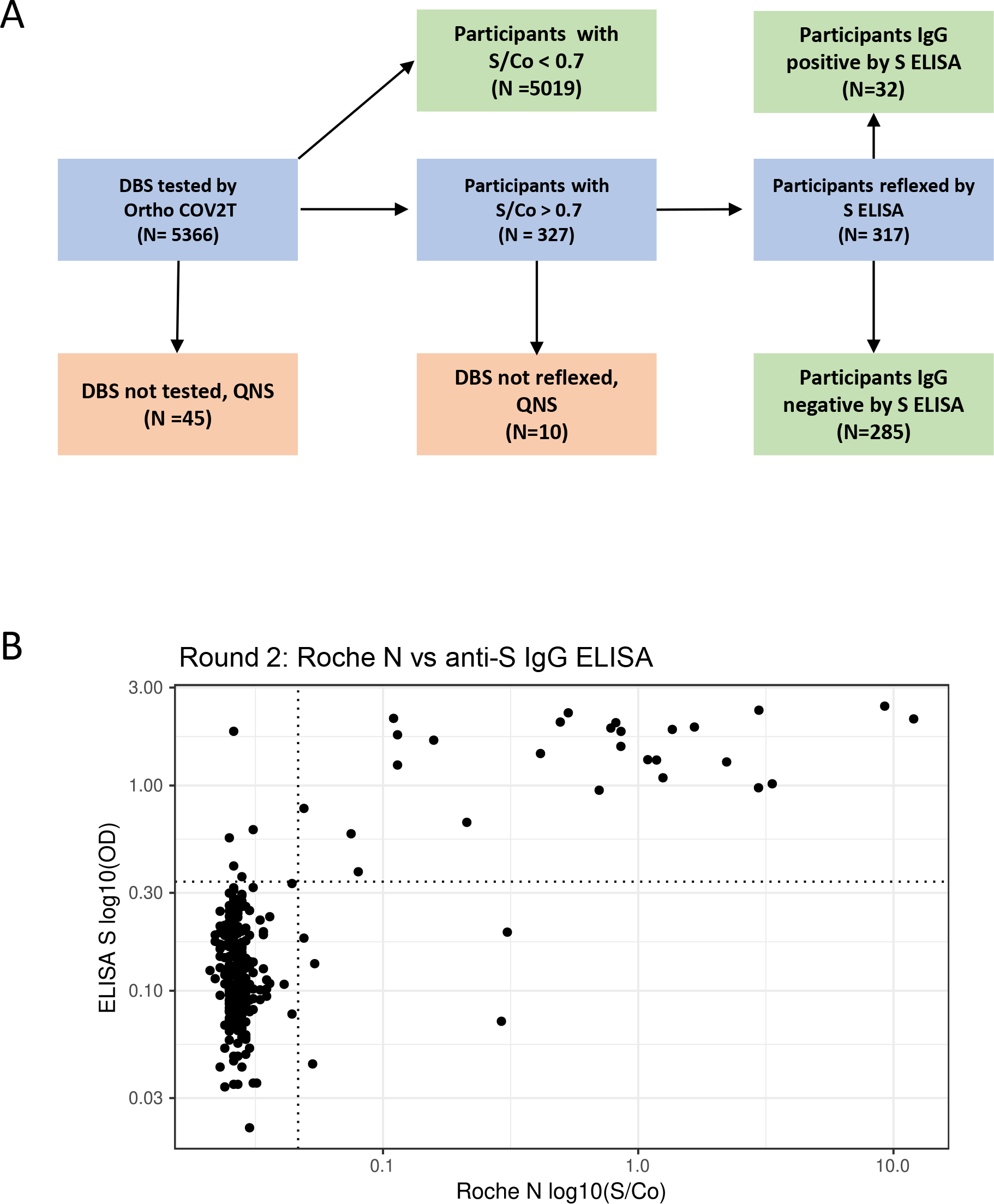
Testing algorithm and results from EBCOVID Round 2. (A) Schematic of the testing algorithm used for Round 2 of the EBCOVID study. QNS=Quality not sufficient. (B) Round 2 EBCOVID results comparing DBS reflexed according to the testing algorithm on the anti-S IgG ELISA and the Roche N assays.

During Round 3 (February to April 2021), vaccines became available to the public, including our study participants. To distinguish between natural infection and vaccine-induced antibody responses, we modified our testing algorithm to include reflex testing for anti-N antibody using the Roche N assay. Round 3 samples were tested first on the Ortho CoV2T assay, and all anti-S-antibody reactive samples with an S/Co ratio >0.7 and <2.0 were reflexed to the anti-S IgG ELISA, while all anti-S-antibody samples with an S/Co >0.7 were reflexed to the Roche N DBS assay. We considered a confirmed natural infection as any sample that was positive by anti-S IgG ELISA or had an Ortho CoV2T Assay S/C ratio greater than 2.0 and was positive in the Roche N DBS assay. Using this algorithm, we determined that, of 4,641 samples tested, 1,452 (31.3%) had anti-S antibodies and 84 (1.8%) had anti-NC antibodies due to natural infection (Figure 5).

**Figure 5.**
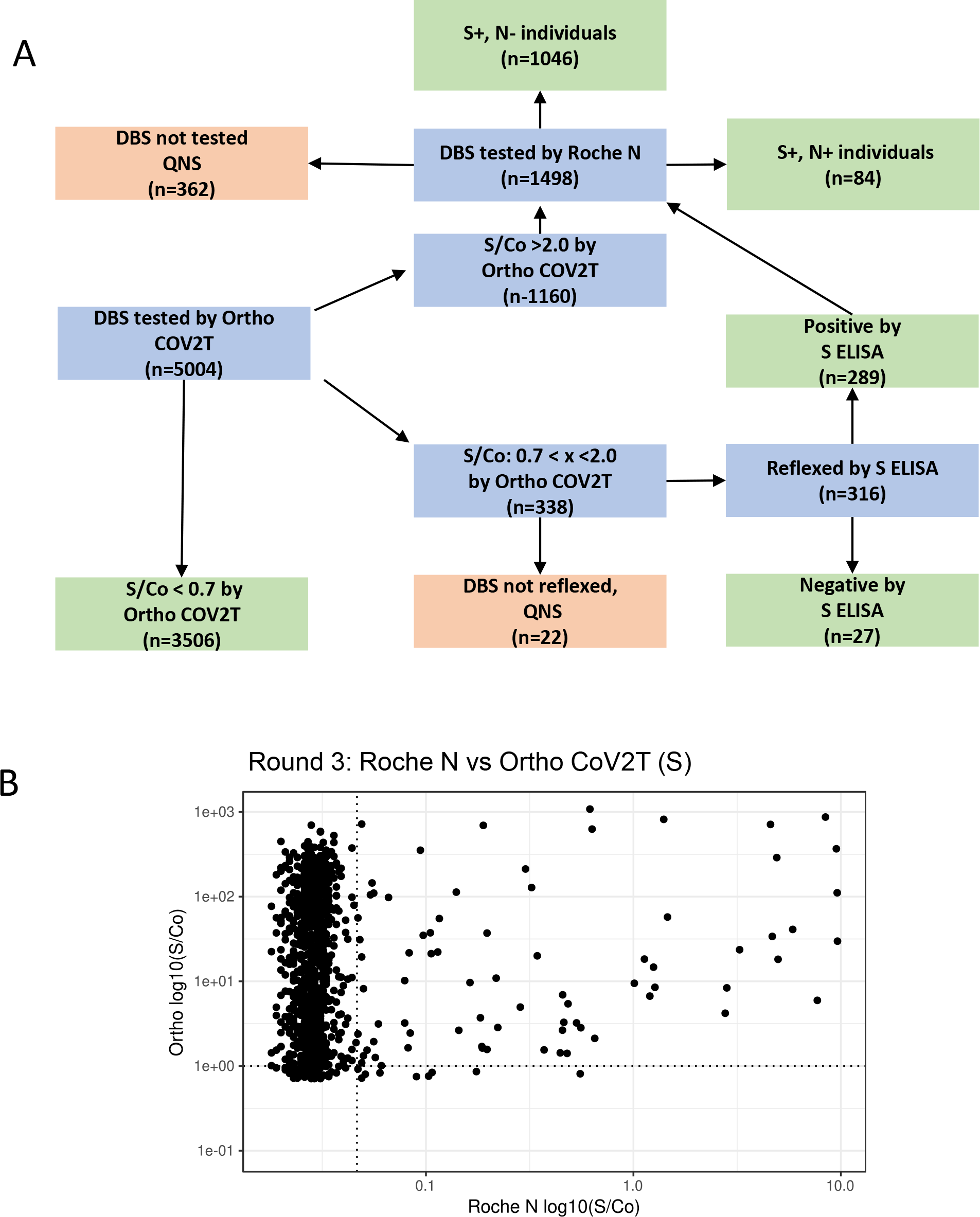
Testing algorithm and results from EBCOVID Round 3. (A) Schematic of the testing algorithm used for Round 3 of the EBCOVID study. QNS=Quality not sufficient. (B) Round 3 EBCOVID results comparing DBS reflexed according to the testing algorithm on the Ortho CoV2T (Ortho S) and the Roche N assays.

### Anti-S antibodies remain stable over time whereas anti-N antibodies appear to wane

Since a key characteristic needed for our study as well as other serosurveillance studies is the ability to accurately track cumulative incidence of infection, we empirically assessed whether the DBS format affected the ability of our serological assays to detect long-term, persistent antibody responses to S and N. We generated a panel of plasma and reconstituted whole blood DBS from 10 COVID-19 convalescent plasma donors sampled longitudinally between 0 and 246 days from their first donation and tested this panel using our in-house indirect IgG ELISAs for S and N, the Ortho CoV2T (S) assay, and the Roche N Total Ig assay. The Ortho CoV2T assay showed stable antibody reactivity over time in plasma and DBS eluates; however, the DBS eluate signal was substantially lower than the signal from neat plasma samples. Our in-house indirect S IgG ELISA also showed antibody stability in both DBS and plasma formats over time, but with similar reactivity observed for plasma and DBS eluates (Figure 6A). In contrast, both the Roche N Total Ig and our in-house indirect N IgG ELISA showed a decrease in antibody reactivity over time in 6 of 10 donors tested (Figure 6B), although every donor would have still been considered antibody reactive. The signal magnitude of the in-house N ELISA, in both DBS and plasma formats, were comparable to each other as well as to the Roche N plasma results, whereas the Roche N DBS results displayed an overall reduction in signal (Figure 6B). Overall, we found that our serological assays in both plasma and DBS formats were suitable for detection of long-term antibodies against S and N.

**Figure 6.**
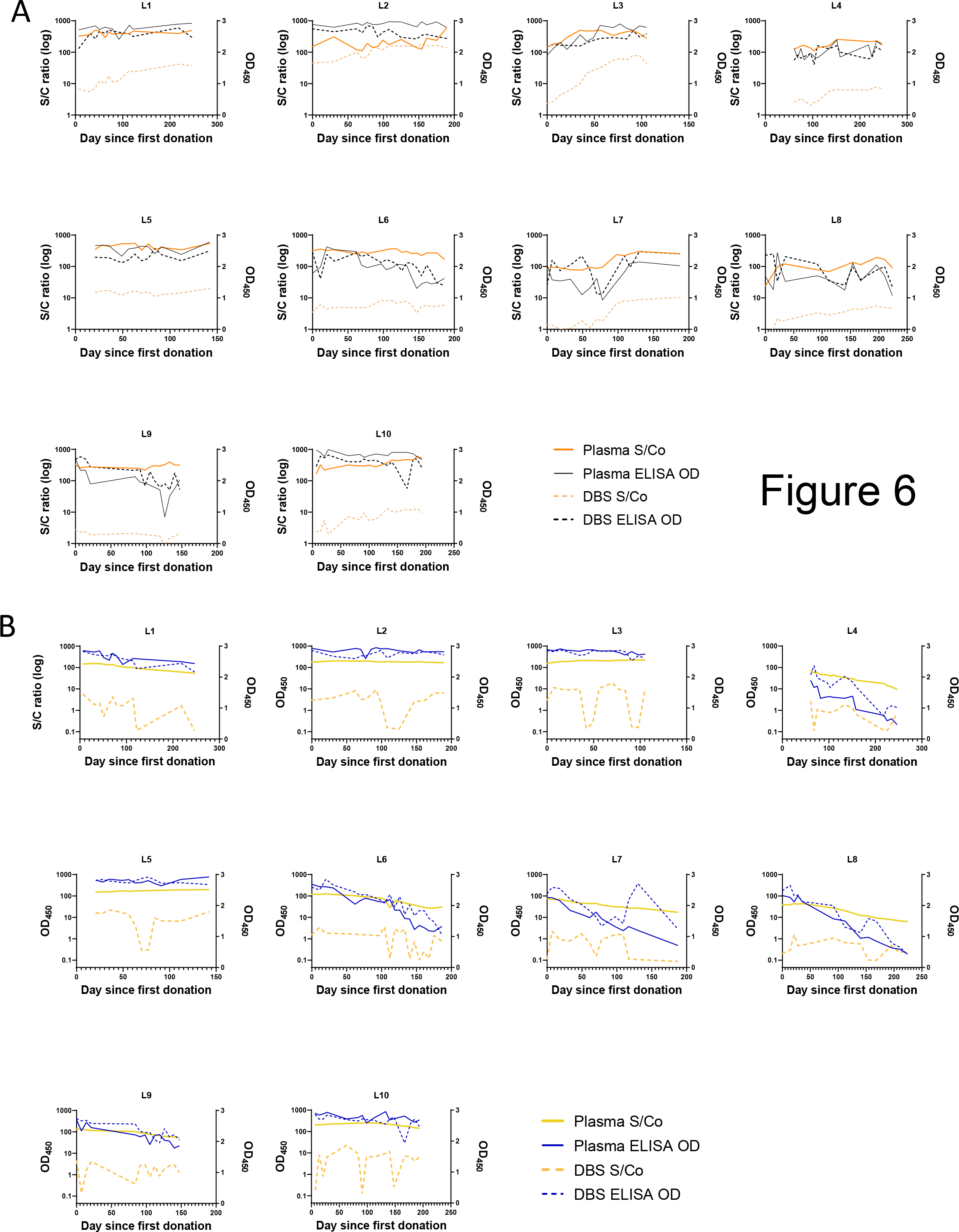
Durable SARS-CoV-2 antibody responses to S and N can be detected by DBS. Paired plasma (solid lines) and DBS samples (dashed lines) from 10 COVID-19 convalescent plasma donors (L1-L10) sampled longitudinally between 0 and 246 days from their first donation were analyzed by (A) anti-S IgG ELISA and the Ortho CoV2T assay and (B) anti-N ELISA and the Roche N assay.

### DBS eluates reflect antibody responses after vaccination

We next evaluated the ability of our assays to measure the antibody response after vaccination against SARS-CoV-2. We generated a longitudinal panel of DBS following 12 vaccinated individuals sampled before their first dose, after their first dose, and after their second dose and tested this panel using the Ortho CoV2T assay and anti-S ELISA. We found that the while both assays were able to capture the increase of antibodies after the first dose of vaccine, the anti-S DBS ELISA did not show subsequent boosting of the antibody response after the second dose when plotted by optical density (OD) (Figure 7A-C). We reasoned that this was due to saturation of the OD of the anti-S-ELISA; therefore, we performed dilutions of the DBS eluate and generated an endpoint titer for each sample. Determining the endpoint titer effectively increased the dynamic range of the anti-S DBS ELISA and allowed us to capture boosting of the antibody response due to the second dose of vaccine (Figure 7D). We further validated this concept by following the vaccine antibody response via weekly DBS sampling of a small cohort of 4 individuals over a period of 6 months. Comparisons between the single-dilution OD reading and the endpoint titers revealed that while the single-dilution OD stayed stable across the sampling period, the endpoint titers showed an increase in titers up to four weeks after the second dose and subsequent decline over the next six months (Figure 7E-F). 2 of the individuals also showed an increase in titers after receiving booster doses. These results suggest that our anti-S DBS ELISA is not only able to qualitatively detect persistent antibodies after infection and vaccination but also able to track antibody kinetics by using an endpoint titer method.

**Figure 7.**
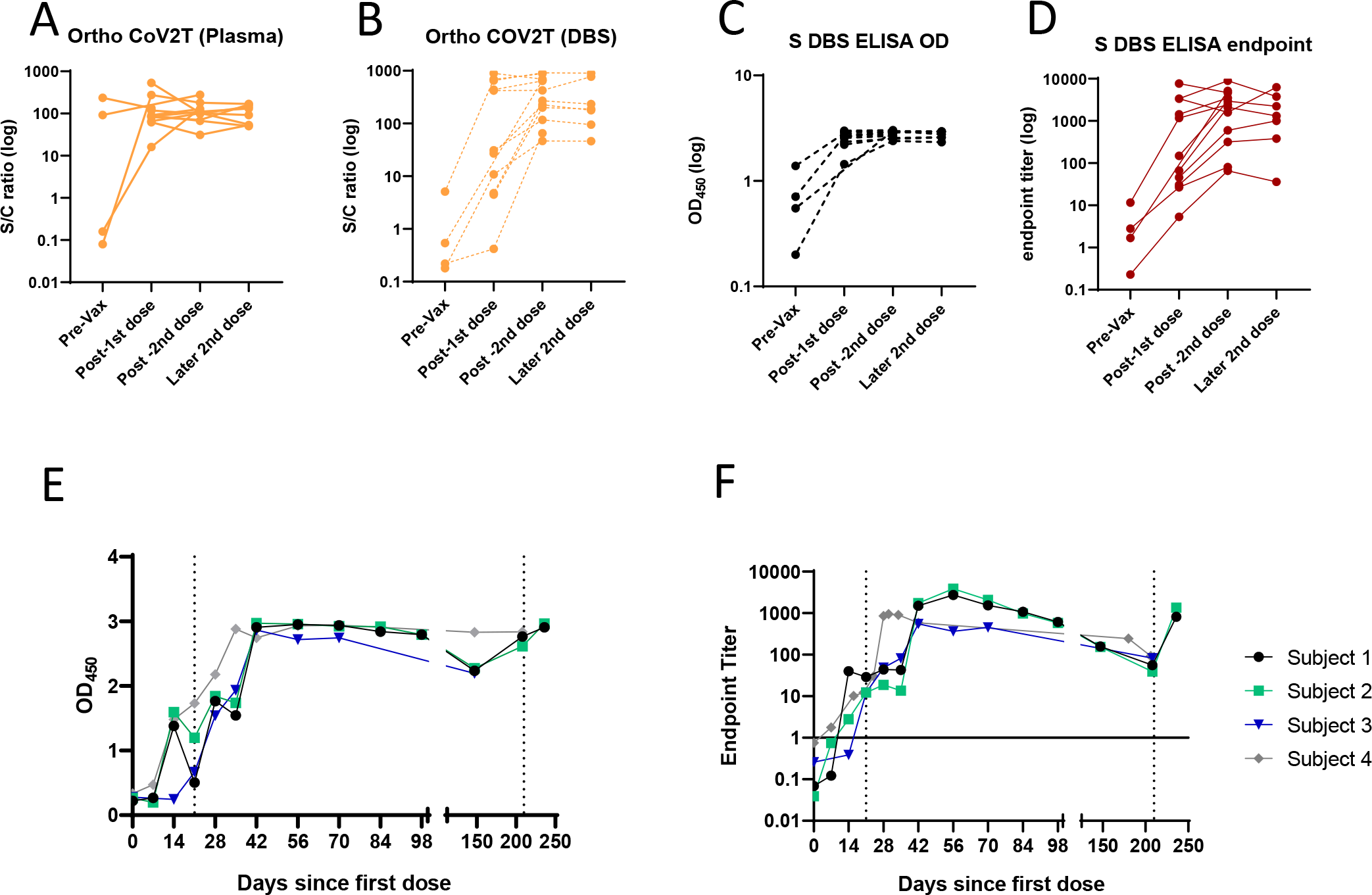
Vaccine-elicited SARS-CoV-2 antibody kinetics can be detected by DBS. (A-B) Plasma and DBS generated from 12 SARS-CoV-2 S-vaccinated individuals sampled before their first dose, after their first dose, and after their second dose were analyzed using the Ortho COV2T assay. DBS from these same individuals were analyzed by the anti-S IgG DBS ELISA as OD_450_ values or (D) endpoint titers. DBS from 4 other vaccinated individuals sampled weekly after their first, second and third doses were analyzed by the anti-S IgG DBS ELISA as (E) OD_450_ value or (F) endpoint titer. Solid line in (F) represents positivity cut-off. Dashed lines in (E-F) denote days where additional doses of vaccine were administered.

## Discussion

This study demonstrates that DBS are a suitable replacement for plasma or serum in serological assays and that fingerstick-derived DBS can be effectively implemented for community-based epidemiologic studies. We validated four distinct DBS serological assays with different assay formats and demonstrated that DBS performed similarly to plasma, can be used to detect antibody responses up to 246 days (or more) after infection, and can be used to track antibody kinetics after vaccination. These assays were implemented in a large longitudinal serological survey of 12 cities in the East Bay region of the San Francisco metropolitan area with at-home biospecimen collection, demonstrating the utility of the DBS format for large-scale serosurveillance.

Recent work on the topic of SARS-CoV-2 DBS serology has shown that DBS samples and serum/plasma samples perform comparably in most serological assay formats (9, 12, 16, 17, 19). One recent study reported that DBS samples could also be adapted for epitope profiling by phage display and for SARS-CoV-2 pseudovirus neutralization (19). These investigators, along with others, validated their serological assays on DBS self-collected by study participants at home and mailed to investigators, providing proof-of-concept that a serological study could be performed using DBS (17, 19, 20). However, these studies analyzed relatively small number of participants, which limited insights into the scalability of this approach for large epidemiologic studies. Several studies adapted the Roche N assay, a high-throughput semi-automated commercial assay, for use on DBS eluates to process a larger number of samples. However, since the Roche N assay only detects antibodies to N, anti-S antibody responses at the population level have yet to be measured in the DBS format. Here, we present extensive validation of both commercial and in-house-derived serological assays against S and N and demonstrate the implementation of these methods in a large longitudinal cohort study involving at-home DBS collection by the study participants.

Our S ELISA showed 100% agreement with plasma and DBS in our validation studies. The Ortho CoV2T assay showed an 80% concordance between DBS and plasma format at the recommended S/Co cutoff of 1.0, displaying somewhat reduced sensitivity. This loss of sensitivity was attributed to the dilutional effect that occurs during the DBS elution process. We determined that our DBS elution method results in an approximately 1:40 dilution compared to plasma, which is in line with other studies that compared total IgG levels between DBS and plasma (19). This dilutional effect explains why ELISAs perform well when adapted to the DBS format; since most indirect ELISAs are performed with a serum/plasma dilution step, they are less affected by any dilution introduced during the DBS elution step. In contrast, the Ortho and Roche Total Ig sandwich assays are performed using undiluted serum/plasma. When optimizing the Roche N assay, we were able to adjust the S/Co threshold from 1 to 0.04 based on our performance data and thus improve the sensitivity of the assay in the DBS format. We compensated for the loss of sensitivity in the Ortho S CoV2T assay by reflexing DBS samples with Ortho CoV2T S/Co values that fell within the range of 0.7 to 2 to the in-house anti-S ELISA.

In this study, we were interested in understanding the prevalence of SARS-CoV-2 infection in our study population. Since we used seropositivity as a marker for past infection, it was important to evaluate whether serological assays in the DBS format would be able to detect durable antibody responses months after initial infection. Studies have shown that the durability of SARS-CoV-2 antibody responses are variable and depend on assay format, target antigens and severity of disease (20–22). While most individuals had persistent detectable antibodies over 42 weeks post-infection, other individuals demonstrated waning of the antibody response, which correlated with being male, older, and/or having milder disease (21). One study comparing SARS-CoV-2 serological assays using a set of COVID-19 convalescent plasma from a longitudinal cohort 63-129 days following resolution of symptoms showed that some assays, like the Ortho COV2T and the Roche N assays, show stable antibody reactivity over time. while others showed a decline in reactivity (23). We assessed the longitudinal performance of all four serological assays employed in our study using a longitudinal sample set derived from 10 SARS-CoV-2 infected convalescent plasma donors with intervals between resolution of disease symptoms and last donation up to 246 days, using both reconstituted whole blood derived DBS and matched plasma samples. We found that all our assays were consistently able to detect a stable antibody response in previously SARS-CoV-2-infected individuals, although N antibody responses did show some waning.

Recent interest has developed in the waning of SARS-CoV-2 antibody responses after vaccination (24–26). Studies have shown that the antibody response induced by the commonly used mRNA vaccine BNT162b2 (Pfizer-BioNTech) shows a peak in antibody titers several weeks after the administration of the second dose with subsequent decline in total antibody levels and neutralizing antibody titers over a 6-month period (24, 25). We showed that an anti-S DBS ELISA could accurately detect these kinetics by performing an endpoint titer on the DBS eluate, adding greater utility to the DBS format. Thus, DBS samples for SARS-CoV-2 can provide valuable information about both seroprevalence at the population level and antibody titers over time at the individual level.

Finally, we have demonstrated the feasibility of using DBS samples in a large longitudinal study. At-home sample collection and the implementation of DBS enabled the collection of valuable serological data from our study population to investigate changes in seroprevalence over three time-points. Additional examples of how DBS serology data derived from large samples can be successfully used to conduct epidemiologic investigations include our own recent studies. We combined DBS results with comprehensive questionnaire and other publicly available data to estimate population-adjusted SARS-CoV-2 seroprevalence and differences by age, sex, race/ethnicity, zip code, and other demographic strata, and to characterize mitigation behaviors and their effects on SARS-CoV-2 seroprevalence (28). In addition, DBS serology results as described here were also used to assess the relationship between vaccination antibody response and several study participant characteristics including age, sex, vaccine type, vaccine side effects and other health-related factors (O. Solomon and L. Barcellos, unpublished data).

In summary, the use of DBS has gained recent interest as an alternative to venous blood sampling due to the COVID-19 pandemic. Many of the strengths of the DBS format (stability at room temperature, less sample volume required, cost-effectiveness) that make them well-suited for serological studies in resource-limited settings also make them an attractive option for SARS-CoV-2 serological surveillance since they enable at-home sampling from study participants. Samples can be returned by mail, safeguarding the health of study participants by avoiding clinic-based phlebotomy. While DBS has been an attractive option for biological sampling in many fields, lack of validation of DBS on commercial assays/platforms and limited examples of successful implementation have led to reluctance in implementation of DBS sampling for serosurveillance (7, 27). Our study presents the validation of two commercial assays, highlighting potential pitfalls due to the dilute nature of the DBS eluate. We also present solutions to mitigate the subsequent reduction in sensitivity when switching from plasma/serum to DBS. We, and others, found that ELISAs were easily modified to accommodate DBS eluates with minimal optimization and with equivalent sensitivity and specificity to plasma/serum. Because ELISAs are also relatively low-cost compared to commercial diagnostic assays, a DBS-based ELISA assay for serosurveillance would be advantageous in resource-limited settings. In addition to providing diagnostic utility, our work and that of others show that DBS eluates can also be used to track antibody kinetics, and can be employed in epitope mapping studies, and even in neutralization assays (19). Our work, combined with evidence from others, makes a strong case for the implementation of DBS, not just for SARS-CoV-2 serological studies but in any other setting where phlebotomy would be impractical.

## Data Availability

All data produced in the present study are available upon reasonable request to the authors

## Acknowledgments

We would like to thank John Pak and Aubree Gordon for the provision of S and N proteins used in the study. We also thank Patrick Hsu and Spotlight Therapeutics for donation of S protein used for assay development. This study was supported by Open Philanthropy Projects, Fast Track grants from Emergent Ventures, the UC Berkeley Innovative Genomics Institute, and the UC Berkeley School of Public Health Center for Population Health.

