## Supplemental for "Development and Implementation of Dried Blood Spot-based COVID-19 Serological Assays for Epidemiologic Studies"

Figure S1

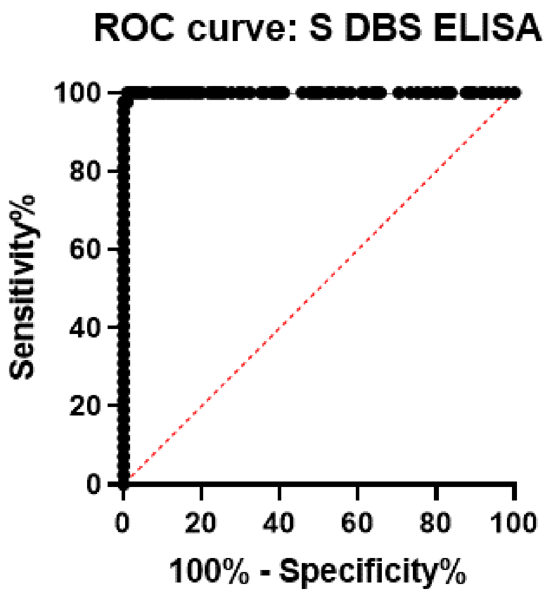


**Supplemental Figure 1.** Receiver-Operator Curve (ROC) of the validation of the anti-S IgG DBS ELISA.

Figure S2

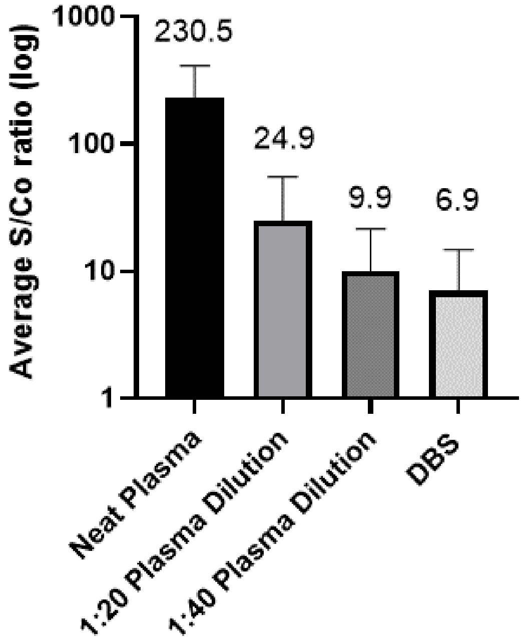


**Supplemental Figure 2.** Comparison of diluted plasma to DBS on the Ortho COV2T assay. Paired plasma-DBS samples (n=37) were tested undiluted or at the indicated dilutions of plasma. Numbers indicate the average S/Co ratio; error bars represent standard deviation.

Figure S3

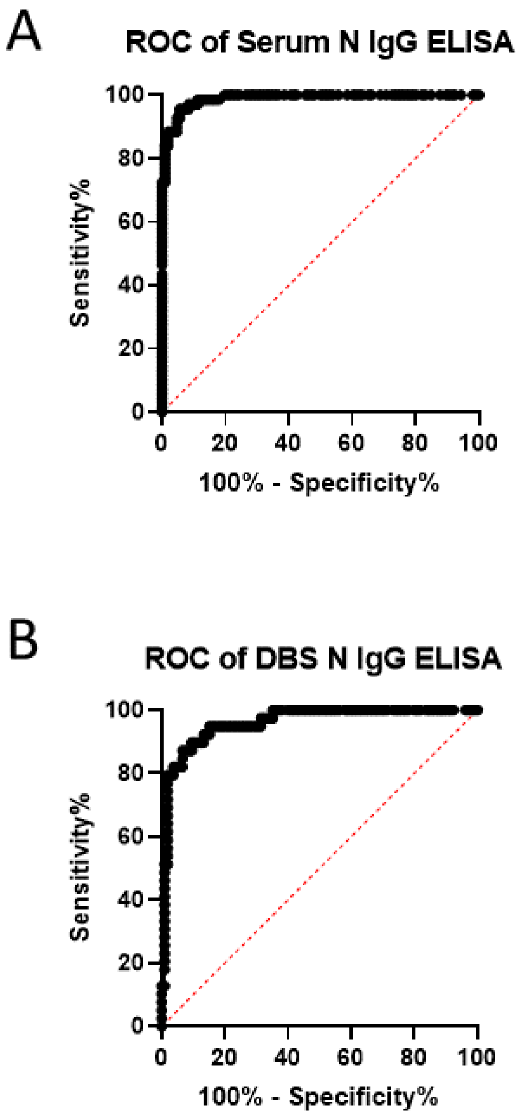


**Supplemental Figure 3.** Receiver-Operator Curves (ROC) of the validation of (A) the anti-N IgG ELISA in serum or of (B) the anti-N DBS ELISA.

Figure S4

A

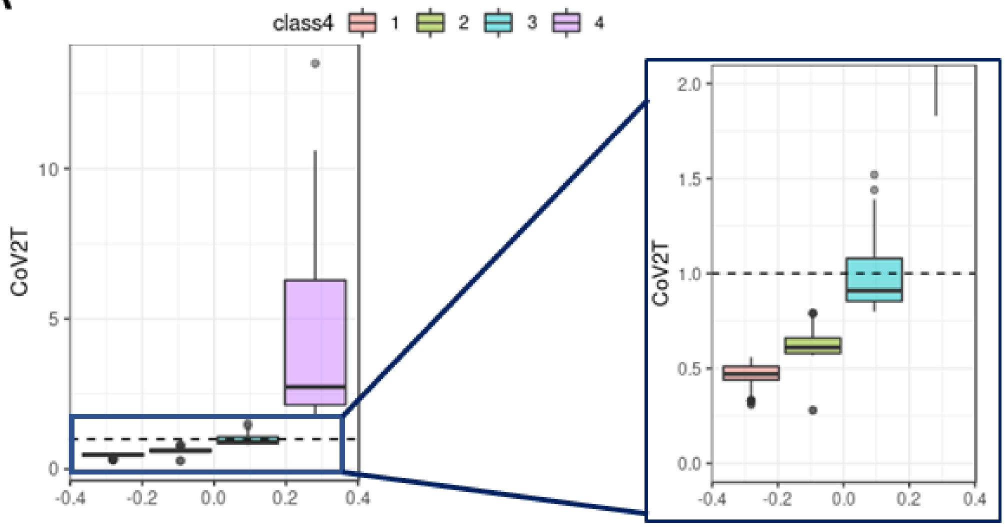


B

| | N | % of Total | Mean S/Co | SD |
|---|---|---|---|---|
| Class 1 | 3753 | 85.88% | 0.472 | 0.0471 |
| Class 2 | 559 | 12.79% | 0.626 | 0.0586 |
| Class 3 | 47 | 1.08% | 0.997 | 0.195 |
| Class 4 | 11 | 0.25% | 4.87 | 4.08 |
| Total | 4370 | 100% | | |

**Supplemental Figure 4.** (A) Ortho COV2T results from Round 1 of the EBCOVID study grouped into 4 categories. Right panel is an inset of the boxed area in the left panel. Dotted line represents the positivity cut-off value. (B) Descriptive table of data in (A). Highlighted row refers to results that fall within the indeterminate zone.
